# External Validation of a Mobile Clinical Decision Support System for Diarrhea Etiology Prediction in Children: A Multicenter Study in Bangladesh and Mali

**DOI:** 10.1101/2021.07.31.21261145

**Authors:** Stephanie C Garbern, Eric J Nelson, Sabiha Nasrin, Adama Mamby Keita, Ben J Brintz, Monique Gainey, Henry Badji, Dilruba Nasrin, Joel Howard, Mami Taniuchi, James A. Platts-Mills, Karen L Kotloff, Rashidul Haque, Adam C Levine, Samba O Sow, Nur H Alam, Daniel T Leung

**Affiliations:** Department of Emergency Medicine, Alpert Medical School, Brown University, Providence, RI, USA; Departments of Pediatrics and Environmental and Global Health, Emerging Pathogens Institute, University of Florida, Gainesville, FL, USA; International Centre for Diarrhoeal Disease Research, Bangladesh (icddr,b), Dhaka, Bangladesh; Center for Vaccine Development - Mali, Bamako, Mali; Division of Epidemiology, University of Utah, Salt Lake City, UT, USA; Rhode Island Hospital, Providence, RI, USA; Center for Vaccine Development and Global Health, University of Maryland School of Medicine, Baltimore, MD, USA; Department, University of Kentucky, Lexington, KY, USA; Division of Infectious Diseases and International Health, University of Virginia, USA; *Department, University of Maryland, Baltimore, MD, USA; Division of Infectious Diseases, University of Utah School of Medicine, Salt Lake City, Utah, USA

**Keywords:** diarrhea, diarrhoea, antibiotic stewardship, antimicrobial resistance, AMR, enteropathogens, enteric infections, Bangladesh, Mali, mobile health, smartphone, clinical prediction model, clinical decision support

## Abstract

**Background:** Diarrheal illness is a leading cause of antibiotic use for children in low- and middle-income countries. Determination of diarrhea etiology at the point-of-care without reliance on laboratory testing has the potential to reduce inappropriate antibiotic use.

**Methods:** This prospective observational study aimed to develop and externally validate the accuracy of a mobile software application (“App”) for the prediction of viral-only etiology of acute diarrhea in children 0-59 months in Bangladesh and Mali. The App used previously derived and internally validated models using combinations of “patient-intrinsic” information (age, blood in stool, vomiting, breastfeeding status, and mid-upper arm circumference), pre-test odds using location-specific historical prevalence and recent patients, climate, and viral seasonality. Diarrhea etiology was determined with TaqMan Array Card using episode-specific attributable fraction (AFe) >0.5.

**Results:** Of 302 children with acute diarrhea enrolled, 199 had etiologies above the AFe threshold. Viral-only pathogens were detected in 22% of patients in Mali and 63% in Bangladesh. Rotavirus was the most common pathogen detected (16% Mali; 60% Bangladesh). The viral seasonality model had an AUC of 0.754 (0.665-0.843) for the sites combined, with calibration-in-the-large α=-0.393 (−0.455 – -0.331) and calibration slope β=1.287 (1.207 – 1.367). By site, the pre-test odds model performed best in Mali with an AUC of 0.783 (0.705 - 0.86); the viral seasonality model performed best in Bangladesh with AUC 0.710 (0.595 - 0.825).

**Conclusion:** The App accurately identified children with high likelihood of viral-only diarrhea etiology. Further studies to evaluate the App’s potential use in diagnostic and antimicrobial stewardship are underway.

## Introduction

Diarrheal diseases remain a leading cause of morbidity and mortality in children younger than five years worldwide, with approximately one billion episodes and 500,000 deaths annually.^1,2^ While a significant problem in all countries, the greatest burden of pediatric diarrhea exists in low- and middle-income countries (LMICs), primarily in South Asia and sub-Saharan Africa.^2^ Although the majority of diarrhea episodes are self-limiting and the mainstay of diarrhea treatment is rehydration, clinicians must also make decisions regarding appropriate use of diagnostics and for antibiotic prescribing. Guidelines from the World Health Organization (WHO) recommend against antibiotic use for the treatment of pediatric diarrhea, except for specific presentations of diarrhea such as suspicion of *Vibrio cholerae (V. cholerae)* with severe dehydration, blood in stool, or concurrent illness such as severe malnutrition.^3^ For the majority of diarrhea etiologies, antibiotics are not recommended, particularly for viral causes of diarrhea in which antibiotics have no benefit.^4^ Viral pathogens such as rotavirus, sapovirus, and adenovirus, are among the top causes of diarrhea in young children in LMICs, as shown in two large multi-center studies from LMICs, the Global Enteric Multicenter Study (GEMS) and the Malnutrition and Enteric Disease (MAL-ED) study.^5,6^

Laboratory testing by culture or molecular assays are often impractical when treating children with diarrhea in the majority of LMIC clinical settings due to time and resource constraints.^7^ As a result, clinicians often make decisions regarding antibiotic use based on non-evidence based assumptions or broad syndromic guidelines.^8^ Unfortunately, physician judgment has been shown to poorly predict etiology and need for antibiotics in diarrheal infections. For example, patients presenting to Kenyan hospitals with diarrhea showed that syndrome-based guidelines for *Shigella* infection led to the failure to diagnose shigellosis in nearly 90% of cases.^9^ Accurate and cost-effective tools to better determine diarrhea etiology at the point-of-care without relying on laboratory tests are greatly needed to reduce antibiotic overuse while conserving scarce healthcare resources.

Electronic clinical decision support systems (CDSS) incorporating prediction models may offer a solution to the challenges of determining diarrhea etiology in low-resource settings. CDSSs have been used in high-income country (HIC) settings to improve the accuracy of diagnosis and reduce costs by avoiding unnecessary diagnostic tests at the point-of-care.^10^ CDSSs, especially as mHealth applications on smartphone mobile devices, hold great potential for implementing sophisticated clinical prediction models that would otherwise be impossible for providers to calculate manually. These tools can also enable flexibility by clinician choice or automation to optimize the clinical algorithm based on epidemiologic and clinical factors dominant in a given location. Despite opportunities to improve clinical care in a cost-aware mindset, there remains a paucity of data on the use of CDSS for infectious disease etiology determination and to support appropriate antibiotic use in LMICs.^11,12^

Our team previously derived and internally validated a series of clinical prediction models using data from GEMS, integrating characteristics of the current patient’s diarrhea episode (“patient-intrinsic” factors including age, blood in stool, vomiting, breastfeeding status, and mid-upper arm circumference) with external (“patient-extrinsic”) data sources (such as characteristics of recent patients, historical prevalence, climate, and seasonal patterns of viral diarrhea “viral seasonality”) in a modular approach.^13^ The best-performing model (which used patient-intrinsic factors + viral seasonality data) had an internally validated area under the receiver-operating characteristic curve (AUC) of 0.83.^13^ The objective of this study was to prospectively externally validate the models for the prediction of viral-only etiology of diarrhea in children aged 0-59 months in Bangladesh and Mali and demonstrate a proof-of concept for the incorporation of the patient-intrinsic + viral seasonality model into a mobile CDSS software application (“App”) for use in LMIC settings with high diarrheal disease burden.

## Methods

### Study Design and Setting

A prospective, observational cohort study was conducted in Dhaka, Bangladesh and Bamako, Mali. Enrollment was conducted in Bangladesh at the Dhaka Hospital of the International Center for Diarrhoeal Disease Research, Bangladesh (icddr,b) rehydration (short stay) unit and in Mali at the Centres de Santé de Référence (CSREF) and the Centres de Santé Communautaires (CSCOM) in Commune V and VI in Bamako, Mali. These locations were selected because of their geographic proximity to GEMS study sites from which the clinical prediction models were trained, without using the same sites. Participants were enrolled in Bangladesh during November and December 2019 and Mali during January and February 2020. The Dhaka Hospital of the icddr,b provides free clinical services to the population of the capital city of Dhaka, Bangladesh and surrounding rural districts and cares for over 100,000 patients with acute diarrhea each year. The CSREF and CSCOM of communes V and VI in Mali serve a catchment area of 2 million people. CSCOM provides basic care such as family planning, vaccination and outpatients visits, while patients with severe illness are referred to the CSREF where there is capacity for hospital admission for medical conditions and for basic and intermediate surgeries.

### Study Participants and Inclusion/Exclusion Criteria

Patients under five years of age (0-59 months) with symptoms of acute diarrhea were eligible for enrollment. Acute diarrhea was defined as three or more loose stools per day for less than seven days. Patients were excluded using the following criteria: no parent or primary caretaker available for consent, diarrhea lasting seven days or longer, fewer than three loose stools in the prior 24 hours, or having a diagnosis of severe pneumonia, severe sepsis, meningitis, or other condition aside from gastroenteritis.

### Staff Training and Oversight

General practice nurses were hired specifically to collect data at both study sites, and study nurses had no other patient care responsibilities during the study period. Nurses received training in study procedures under the guidance of the research investigators. Training topics included: screening procedures, obtaining informed consent, collecting clinical data and laboratory procedures. Study nurses also received practical hands-on training regarding the use of the App to ensure all nurses were comfortable with entering data and using the devices during clinical workflow.

### Technology Development

Software architecture platform and user interface concepts for the App were derived from two prior related studies.^14,15^ The design herein was intended to demonstrate a ‘live’ proof-of-concept that the models could be successfully configured as a CDSS on a mobile device for high-volume clinical settings in an LMIC. On the input page, clinical variables were restricted to those required for the App (Figure 1). Text was favored over symbology to increase clarity based on prior experience. Computations were initiated locally on the device after pressing ‘calculate’. On the output page, the patient code and randomized calculation code were used to enable joining the digital record to the paper case report form. The design objective was to identify weaknesses in the UI for data entry using paper data entry as the reference standard, and address weaknesses after the study. The code-base was allowed to be iterated once at the transition between the Bangladesh and Mali phases of the study to address engineering challenges exposed during ‘live’ deployment. The probability of a viral-alone etiology was provided, and the probabilities by model type were accessible via a drop-down menu. Data were encrypted for security on the device and upon transfer to a HIPAA compliant server. The code bases consisted of Python (server), Java (web portal interface), Android operating system, and TensorFlor Lite (TFLite) on the Android device. TFLite is an open-source cross-platform deep-learning framework. TFLite converts pre-trained models from Python in TensorFlow to a format that is optimized for speed to run models and store data on mobile devices. The App drew only from patient-intrinsic and viral seasonality models/data sources; climate and prior/recent patient data sources were not used in this study because they did not add to the AUC at the GEMS study sites. The App was configured for English (Bangladesh; version ‘2.0_Nov21’) and French (Mali; version ‘2.2.5’).

**Figure 1.**
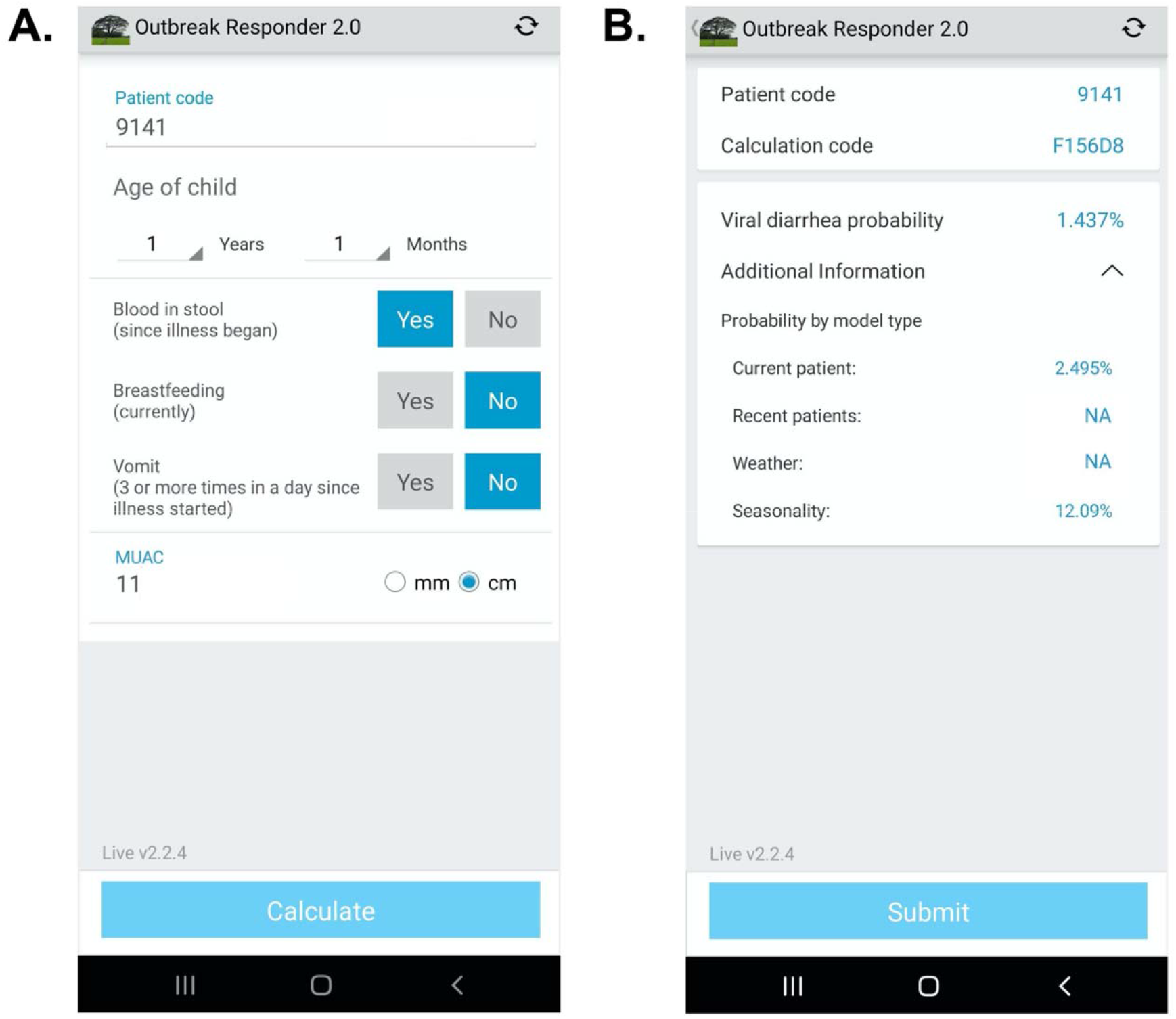
App user interface. **A**. Input page after application launch. **B**. Output page with an example showing calculated probability of viral-only diarrhea. The “^” symbol represents an open accordion menu with the component probabilities. “Current patient” refers to the patient-intrinsic model. “Weather” (climate) and “recent patients” were not active in this configuration.

### Study Procedures

In Bangladesh, due to the high volume of potentially eligible patients presenting daily to icddr,b Dhaka Hospital, study staff randomly selected participants for enrollment on arrival 9am-5pm Sunday to Thursday. Random selection was accomplished using a black pouch filled with white and blue marbles in a preset ratio. Study nurses drew a marble each time a patient presented to the rehydration unit. If a blue marble was pulled, the patient was screened for inclusion and exclusion criteria as described above. After each marble was pulled, it was returned to the bag and shaken. An average of approximately eight patients were enrolled per working day which allowed for high-quality data collection and integrity of all study protocols to be maintained. In Mali, a consecutive sample of patients presenting with acute diarrhea were enrolled. After initial assessment by the facility doctor, children with acute diarrhea were referred to the study team for screening. Study staff were located in the intake area and potential participants’ information was recorded in a screening log. All patients presenting with diarrhea were assessed for eligibility. After screening, research staff provided the parent or guardian with information about the purpose of the study, risks and benefits in Bangla (Bangladesh) and Bambara (Mali) language. Research staff then obtained written consent if the parent or guardian agreed to participate on behalf of the child. In cases where the parent or guardian was illiterate, the consent form was marked with a thumbprint for signature, based on standards for informed consent at icddr,b and CVD-Mali. In these cases, a witness (other than study staff) also signed the consent form. If a child arrived without a parent or guardian in attendance, they were not considered for enrollment in the study.

After enrollment, study staff collected demographic, historical and clinical information from the parent or guardian. All information was collected on a paper case report form (CRF). The ‘patient-intrinsic’ clinical variables were entered into the App on a mobile device (Bangladesh: Samsung Galaxy A51; Mali: Samsung Galaxy Note 10) by two different study nurses independently to ensure reliability; patient-intrinsic variables were age in months for 0-23 months and in years for 2-4 years, blood in stool since illness began, history of vomiting (3 or more times a day since illness started), breastfeeding status (‘currently’), and mid-upper arm circumference (MUAC). During data collection, study investigators noted that nearly all participants in Bangladesh had reported “yes” to the question regarding history of vomiting. Given vomiting was not expected in all patients especially those with non-viral diarrhea etiology, it was determined after speaking with the study nurses that the phrasing of the question sometimes led patients to respond “yes” if there had been regurgitation with feeding or ORS administration, rather than actual vomiting. The question format was then revised for the remainder of the study enrollment in Bangladesh, and prior to any patient enrollment in Mali, to clarify the definition of vomiting. The App calculated the probability of viral diarrhea alone and the probabilities specific to each data source (patient-intrinsic, prior/recent patient, climate and viral seasonality); patient and calculation codes were used to match paper CRFs.

The App results were not used for clinical decision-making to allow first for the external validation and second to iterate the software in response to engineering challenges exposed from ‘live deployment’. All patients were treated according to standard local clinical protocols, and the clinicians caring for patients were blinded to any study data collected in order to prevent any undue influence in clinical care. Study procedures were not allowed to delay any immediately necessary patient care, such as the placement of an intravenous line or delivery of intravenous fluid to the patient.

### Sample Collection and Laboratory Procedures

The first available stool specimen after enrollment of the participant was collected. Study participation concluded after a stool sample was obtained. Nurses were unaware of the etiology of diarrhea at the time of clinical assessment as microbiological testing was conducted only after the study period concluded. Stool samples were collected in a sterile plastic container and then transferred to two separated 2 mL cryovials – one vial with 1 mL stool only and one vial for storage in 70% ethanol (Bangladesh) or 95% ethanol (Mali). Samples were stored at -20C or - 80C freezer for processing. At the conclusion of the study, samples were thawed, underwent bead beating and nucleic acid extraction using the QIAamp Fast DNA Stool Mini Kit. Total Nucleic acid was mixed with PCR buffer and enzyme and loaded onto custom multiplex TaqMan Array Cards (TAC) containing compartmentalized probe-based quantitative real-time PCR (qPCR) assays for 32 pathogens at the icddr,b or CVD-Mali laboratories (see Supplemental File 1 for full list of pathogen targets).

### Assignment of Diarrhea Etiology

The outcome (dependent) variable was defined as the presence or absence of viral-only etiology. Diarrheal etiology was determined for each patient using qPCR attribution models developed previously by Liu et al.^16^ Viral-only diarrhea was defined as a diarrhea episode with at least one viral pathogen with an episode-specific attributable fraction (AFe) threshold of ≥0.5 and no bacterial or parasitic pathogens with an AFe ≥0.5. This clinically relevant outcome measure was selected because patients with viral-only diarrhea should not receive antibiotics. Other etiologies were defined as having a majority attribution of diarrhea episode by at least one other non-viral pathogen. Patients without an attributable pathogen (unknown final etiology for diarrheal episode) were excluded from this analysis since the cause of the diarrheal episode could not be definitively determined. However, prior studies by this research team have shown that these cases have a similar distribution of viral predictions using a model with presenting patient information as those cases with known etiologies and this study had similar results. (Supplemental File 2).

### Data Processing, Model Development and Internal Validation

Clinical prediction models for the outcome of viral etiology of diarrhea were previously derived and internally validated with full details previously published.^13,17^ Briefly, logistic regression models were derived using clinical, historical, anthropometric and microbiologic data from the GEMS study, a large case-control study conducted at seven sites in Asia and Africa (The Gambia, Kenya, Mali, Mozambique, Bangladesh, India and Pakistan) which enrolled 22,568 children under five years.^5^ Using 5-fold repeated cross-validation, models of various sizes were assessed using AUC, with variable selection based on random forest variable importance measures. A logistic regression model was fit using the top five variables (only variables accessible to clinicians at the point-of-care were considered): age, blood in stool, vomiting, breastfeeding status, and MUAC, an indicator of nutritional status.^13^ Demographics, predictors and viral-only outcome data from the development datasets from GEMS in Bangladesh and Mali are shown in Supplemental File 3.

The post-test odds formulation was used to combine the likelihood ratios derived from these independent models along with pre-test odds into a single prediction. This approach allowed multiple components (models) of the formulation to be flexibly included or excluded depending on availability. The information used for the independently trained models included: patient-intrinsic data, climate data, and local viral diarrhea seasonal curves (“viral seasonality”). Climate data (temperature and rainfall) was included given known weather-related patterns of enteric pathogens such as rotavirus, norovirus and cholera. Local weather data proximate to each site’s health centers was obtained using the National Oceanic and Atmospheric Administration (NOAA) Integrated Surface Database. The information used for pre-test odds include: a daily aggregate of historical patient data, or aggregate predictions from recent patient data (the prior four weeks). In order to externally validate the predictions from the post-test odds, we processed the data from this study to match the variables used in those models as closely as possible.

### External Data - Seasonality, Climate

Standardized seasonal sine and cosine curves were used to model the country-specific seasonal patterns of viral etiology of diarrhea.^13^

Functions 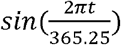 and 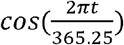, where *t* is based on the date, with a periodicity of 1 year as features to model site-specific seasonality were used. Additionally, climate model features include rain and temperature averages based on a two-week aggregation of the inverse-distance weighted average of the nearest five NOAA-affiliated weather stations to the hospital sites. Weather stations at a distance of greater than 200 km were excluded. The seasonal sine and cosine values as well as temperature and rain averages (climate) were calculated for the dates in this study as described previously.^13^

### Data Analysis

For the purpose of the primary analysis, MUAC measurements collected by the two study nurses were averaged. Patients were considered to have “bloody stool” only if report from both nurses agreed on bloody stool. For children older than two years, age in months was rounded down to the nearest year in months (i.e., 42 months was rounded to 36 months) to match the user interface on the software. Using clinical information gathered from the four data sources (patient-intrinsic, prior patient, climate, viral seasonality), predictions using post-test odds formulation with the developed models were made. The primary model deployed in the App, selected based on the best-performing model from the derivation and internal validation, used the patient-intrinsic data and viral seasonality components.

Model performance for the prediction of viral-only diarrhea was calculated using AUC for each model to evaluate discrimination; calibration was assessed using calibration-in-the-large and calibration slope.^18^ We estimated the calibration coefficients by regressing predicted values versus the observed proportion of viral cases, calculated using the observed proportion of viral cases within 0.05 plus or minus the predicted probability. For the primary analysis, data from the time period using the original vomiting question in Bangladesh was excluded; however, site specific results incorporate all Bangladesh data. The reliability of the predictor data entered independently by the two study nurses was assessed using Cohen’s kappa coefficient (κ) which is a calculation of inter-observer agreement for categorical data.

### Secondary Analysis

The performance of additional models integrating other available components was assessed in post-hoc secondary analysis for comparison. These alternate models included the following components: 1) patient-intrinsic data only 2) patient-intrinsic + climate data (climate) 3) patient-intrinsic + historical patient pre-test odds (historical) 4) patient-intrinsic + prior patient pre-test odds (recent).

For all analyses, a two-tailed p value of 0.05 was considered statistically significant. R (R Foundation for Statistical Computing, Vienna, Austria) were used for all analyses. Standard guidelines from the transparent reporting of a multivariable prediction model for individual diagnosis (TRIPOD) Checklist for Prediction Model Validation were used. The de-identified dataset is available online in Supplemental File 8.

### Sample Size Calculation

The previously derived clinical prediction model had an internally validated AUC of 0.83.^13^ In order to ensure that the confidence intervals around the estimates in the validation study would not cross 0.75 (generally considered to be a marker of adequate accuracy for a clinical prediction model) a margin of error around the AUC of 0.08 was required. Using the approximate variance estimate for AUC from the literature, and assuming a prevalence of a viral only etiology of approximately 30% in our sample, a sample size of 300 patients (150 patients per site) was required.

## Results

### Participant Characteristics

A total of 302 patients were recruited from the two study sites with 152 patients in Bangladesh and 150 patients in Mali. All patients except two in Bangladesh had a stool sample collected for TAC testing, for a total of 300 patients with TAC results (Figure 2). Diarrhea etiology was assigned for a total of 199 patients (66%; 130 in Bangladesh and 69 in Mali) for inclusion in the final analysis (Table 1). The median [IQR] age of included patients was 12 months; there was a predominance of male patients at both study sites (61.8% overall) with 59.2% in Bangladesh and 66.7% in Mali. Sociodemographic and clinical characteristics of the study participants are shown in Table 1. In Bangladesh, 73 patients were asked about vomiting history using the original question format and 57 patients using the revised question format.

**Table 1.** Clinical Characteristics of Study Population.

|  | Diarrhea Etiology Assigned |  |  | No Diarrhea Etiology Assigned |  |  |
| --- | --- | --- | --- | --- | --- | --- |
|  | Overall<br>n(%)<br>N=199 | Bangladesh<br>n(%)<br>N=130 | Mali<br>n(%)<br>N=69 | Overall<br>n(%)<br>N=101 | Bangladesh<br>n(%)<br>N=20 | Mali<br>n(%)<br>N=81 |
| Age (median, IQR),<br>months | 12 (8) | 12 (9) | 11 (8) | 8 (7) | 11.5 (7) | 8 (7) |
| Sex |  |  |  |  |  |  |
| Male | 123 (61.8) | 77 (59.2) | 46 (66.7) | 63 (62.4) | 11 (55) | 52 (64.2) |
| Female | 76 (38.2) | 53 (40.8) | 23 (33.3) | 38 (37.6) | 9 (45) | 29 (35.8) |
| Diarrhea Duration<br>(median, IQR), days | 2 (2) | 3 (1) | 0.6 (0.3) | 1.5 (2.3) | 3 (1.3) | 0.7(0.1) |
| # Episodes of Diarrhea<br>Past 24 hours (median,<br>IQR) | 12 (9.5) | 15 (8) | 5 (3) | 6 (4) | 15 (3.5) | 5 (3) |
| Bloody Stool Reported |  |  |  |  |  |  |
| Yes | 7 (3.5) | 2 (1.5) | 5 (7.2) | 3 (3) | 0 (0) | 3 (3.7) |
| No | 192 (96.5) | 128 (98.5) | 64 (92.8) | 98 (97) | 20 (100) | 78 (96.3) |
| Fever Reported |  |  |  |  |  |  |
| Yes | 163 (81.9) | 119 (91.5) | 44 (63.8) | 66 (65.3) | 18 (90) | 48 (59.3) |
| No | 36 (18.1) | 11 (8.5) | 25 (36.2) | 35 (34.7) | 2 (10) | 33 (40.7) |
| Vomiting Reported** |  |  |  |  |  |  |
| Yes (original question<br>format) | 73 (36.7) | 73 (56.2) |  | 12 (11.9) | 12 (60) |  |
| Yes (revised question<br>format) | 81 (40.7) | 45 (34.6) | 36 (52.2) | 21 (20.8) | 5 (25) | 16 (19.8) |
| No | 45 (22.6) | 12 (9.2) | 33 (47.8) | 68 (67.3) | 3 (15) | 65 (80.2) |
| Breastfeeding |  |  |  |  |  |  |
| Yes (Partial or<br>Exclusive) | 155 (77.9) | 99 (76.2) | 56 (81.2) | 89 (88.1) | 19 (95) | 70 (86.4) |
| No | 44 (22.1) | 31 (23.8) | 13 (18.8) | 12 (11.9) | 1 (5) | 11 (13.6) |
| MUAC (median, IQR),<br>cm | 13.55 (1.4) | 13.45 (1.3) | 13.8 (1.5) | 13.35 (1.8) | 13.375 (2.2) | 13.35 (1.9) |
| Prior Medications Taken |  |  |  |  |  |  |
| Yes | 124 (62.3) | 109 (83.8) | 15 (21.7) | 34 (33.7) | 17 (85) | 17 (21) |
| No | 75 (37.7) | 21 (16.2) | 54 (78.3) | 67 (66.3) | 3 (15) | 64 (79) |
| Years of Mother's<br>Education | 8 (6) | 8 (5) | 4 (9) | 6 (10) | 8 (4.75) | 5 (10) |
| Years of Father's<br>Education | 8 (6) | 8 (5) | 6 (9) | 6 (10) | 10 (4.8) | 4 (10) |
| People living at home<br>(median, IQR) | 6 (4) | 5 (2) | 9 (12) | 9 (10) | 6 (6) | 10 (9) |
| Abbreviations: IQR, interquartile range; cm, centimeter; MUAC, mid-upper arm circumference |  |  |  |  |  |  |
| ** Vomiting question was asked in two different formats at the Bangladesh site |  |  |  |  |  |  |

**Figure 2.**
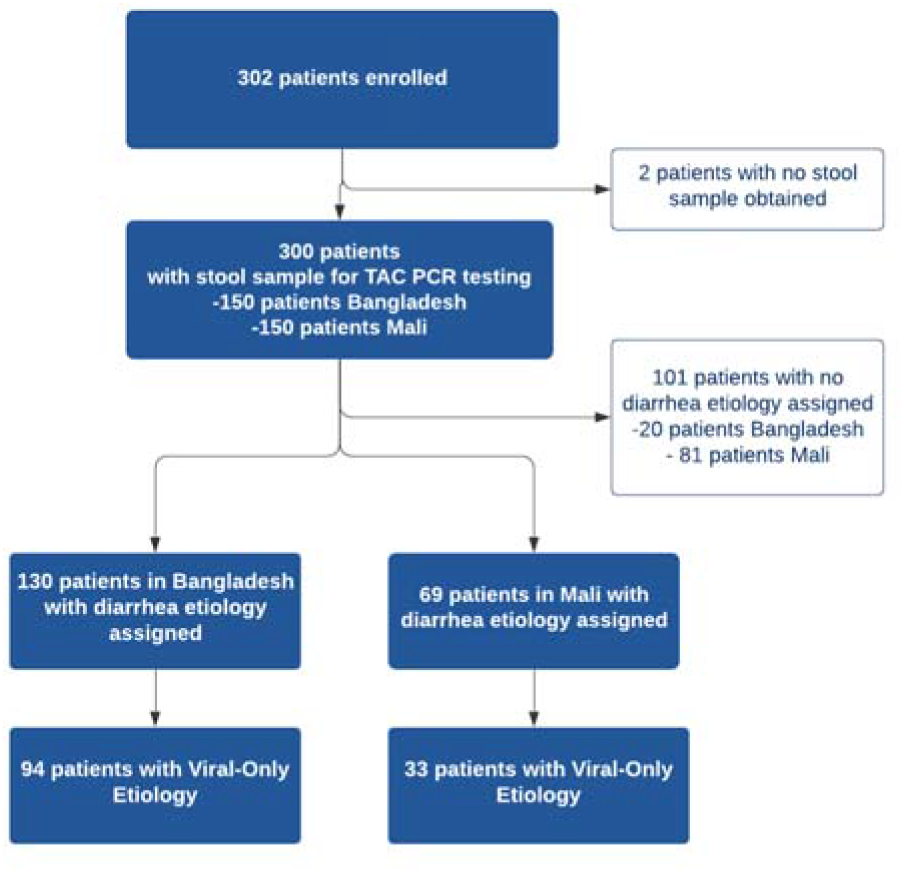
Study Flow Diagram.

### Diarrhea Etiology

Viral-only etiologies of diarrhea were the predominant cause of illness among patients with assigned etiology overall (42%) although viral-only etiologies were more common in Bangladesh (63%) compared to Mali (22.0%). Rotavirus was the most common viral pathogen detected in both study sites (38% overall) while the rate was higher in Bangladesh (60%) compared to Mali (16%). Other viral causes (e.g., adenovirus, astrovirus, norovirus) were less common in both sites. Diarrhea etiology was unable to be assigned in a substantial proportion of the patients in Mali (54%) compared to Bangladesh (13.3%). The prevalence of the various pathogens detected are listed in Table 2.

**Table 2.** Pathogens Detected with TaqMan Array Card by Study Site.

| Bangladesh<br>N=150 |  | Mali<br>N=150 |  |
| --- | --- | --- | --- |
| Pathogen | n (%) | Pathogen | n (%) |
| No Etiology Assigned | 20 (13) | No Etiology Assigned | 81 (54) |
| ***Viral-Only Pathogens*** | 94 (63) | ***Viral-Only Pathogens*** | 33 (22) |
| Rotavirus | 90 (60) | Rotavirus | 24 (16) |
| Adenovirus 40/41 | 1 (0.6) | Norovirus GII | 5 (3.3) |
| Astrovirus | 2 (1.3) | Astrovirus | 2 (1.3) |
| Adenovirus & Astrovirus | 1 (0.6) | Astrovirus & Rotavirus | 2 (1.3) |
| ***Bacterial-Only Pathogens*** | 9 (6) | ***Bacterial-Only Pathogens*** | 24 (16) |
| *Vibrio cholerae* | 3 (2) | *Shigella* / EIEC | 13 (8.7) |
| *Shigella* / Enteroinvasive *E. coli* (EIEC) | 1 (0.6) | Shiga-toxin Enterotoxigenic *E. coli* | 4 (2.7) |
| *Campylobacter jejuni* / *coli* | 1 (0.6) | *Campylobacter jejuni* / *coli* | 2 (1.3) |
| Multiple Bacterial Pathogens | 4 (2.7) | *Salmonella* | 2 (1.3) |
|  |  | Multiple Bacterial Pathogens | 3 (2) |
| ***Parasitic Pathogens*** | 1 (0.6) | ***Parasitic Pathogens*** | 5 (3.3) |
| Cryptosporidium | 1 (0.6) | Cryptosporidium | 4 (2.7) |
|  |  | *Entamoeba histolytica* | 1 (0.6) |
| ***Mixed Pathogens*** | 26 (17) | ***Mixed Pathogens*** | 7 (4.7) |
| Viral + Bacteria | 24 (16) | Viral + Bacteria | 7 (4.7) |
| Viral + Bacterial + Parasitic | 2 (1.3) |  |  |

### Clinical Prediction Models Performance: Primary and Secondary Models

When applied to both sites, the primary model which included patient-intrinsic + viral seasonality component performed better than secondary models at discriminating a viral-only etiology from all other known etiologies with an AUC of 0.754 (95% CI 0.665-0.843) (Table 3). However, this same model was not well calibrated with an estimate of calibration-in-the-large of α=-0.393 (95% CI -0.455 to -0.331) and a calibration slope of β=1.287 (95% CI 1.207, 1.367). The model with pre-test odds calculated using historical prevalence data, while having low discrimination (AUC 0.702 (95%CI 0.603 to 0.800) was the best calibrated with estimates of 0.036 (−0.031 – 0.102) and 1.063 (0.943 – 1.184) for α and β, respectively. Combinations of more than two components including the patient-intrinsic information reduced the discriminatory performance of the model, likely due the positive correlation between components.

**Table 3.** Model performance using AUC (95% confidence interval), calibration-in-the-large (α), calibration slope (β) for each model considered at both sites. Each row after “Patient-Intrinsic Only” includes the Patient-Intrinsic component.

| | AUC (95% CI) | $\alpha$ | $\beta$ |
| --- | --- | --- | --- |
| **Patient-Intrinsic Only** | 0.744 (0.651-0.836) | -0.212 (-0.264 - -0.16) | 1.250 (1.171 - 1.329) |
| **Viral Seasonality** | 0.754 (0.665-0.843) | -0.393 (-0.455 - -0.331) | 1.287 (1.207 - 1.367) |
| **Climate** | 0.731 (0.638-0.825) | -0.255 (-0.292 - -0.218) | 1.140 (1.091 - 1.188) |
| **Historical Patient** | 0.702 (0.603-0.800) | 0.036 (-0.031 - 0.102) | 1.063 (0.943 - 1.184) |
| **Recent Patient** | 0.737 (0.671-0.793) | -0.253 (-0.287 - -0.22) | 1.165 (1.12 - 1.21) |

When looking at the sites individually, the model with pre-test odds calculated using recent patient data and patient-intrinsic data performed best in Mali with an AUC of 0.783 (95% CI 0.705 - 0.86) while the primary model with the viral seasonality component and patient-intrinsic data performed best in Bangladesh with an AUC of 0.71 (95% CI 0.595 - 0.825) (Table 4). When the dates where the vomiting question was asked incorrectly are included in the analysis, all models performed less well with the viral seasonality and climate models performing best in Bangladesh with an AUC of 0.610 (95% CI: 0.523 – 0.697) and 0.617 (95% CI: 0.538 – 0.696),respectively (Table 4).

**Table 4.** AUC (95% confidence interval) for each model by site. Each row after “Patient-Intrinsic Only” includes the Patient-Intrinsic component. The last column includes Bangladesh data in which the vomiting question was asked incorrectly.

|  | Mali | Bangladesh | Bangladesh (no date restriction) |
| --- | --- | --- | --- |
| **Patient-Intrinsic** | 0.763 (0.681 - 0.844) | 0.692 (0.572 - 0.812) | 0.607 (0.521-0.693) |
| **Viral Seasonality** | 0.742 (0.659 - 0.825) | 0.71 (0.595 - 0.825) | 0.61 (0.523 - 0.697) |
| **Climate** | 0.717 (0.63 - 0.804) | 0.679 (0.56 - 0.799) | 0.617 (0.538 - 0.696) |
| **Historical Patient** | 0.741 (0.658 - 0.824) | 0.646 (0.516 - 0.775) | 0.592 (0.505 - 0.86) |
| **Recent Patient** | 0.783 (0.705 - 0.86) | 0.625 (0.5 - 0.75) | 0.61 (0.526 - 0.694) |

### Reliability of Software Use

The agreement between the two study nurses’ independent assessments of the required predictor variables and estimated viral-only etiology risk calculated by the App for each nurse’s assessment was evaluated and showed excellent reliability and agreement between the nurses; results are shown in Supplemental File 4. Data from the App were recorded through the cellular network to a database as well as on the paper CRF. To monitor integrity of the data transfer to the App database from a technical perspective, data were compared between the two records. The agreement between the App database and study nurse’s paper CRF are shown in Supplemental Files 5-6. The Bangladesh study exposed that one field (‘breastfeeding’) did not sync successfully which was addressed at the midpoint between the Bangladesh and Mali study phases. The Mali study used an updated software version (v2.2.5); the reliability and agreement between the App prediction of viral etiology with the post-hoc predictions are shown in Supplemental File 7.

## Discussion

This prospective, clinical study provides an external validation of a mobile CDSS for the prediction of diarrhea etiology in children under five years old. Combining the strengths of prediction models generated through machine learning, the novelty of etiological prediction, and the accessibility gained by incorporation into software on a mobile device, this study represents an important advancement in translational research to build evidence-based tools to improving care in resource-limited settings. Incorporating the internally and now externally validated etiology prediction tool as a component of CDSS for diarrheal diseases could have broad scalable impact, including the potential for reducing inappropriate antibiotic use.

Despite recommendations from the WHO discouraging antibiotic use for the majority of presentations of pediatric diarrhea, multiple studies from LMICs have shown that antibiotic overuse and inappropriate antibiotic prescribing remains widespread with up to 50-80% of children with diarrhea under five receiving antibiotics.^7,19–22^ While antibiotic use is widespread in HICs and LMICs, antibiotic overuse is particularly challenging in LMICs for a multitude of reasons including: widespread availability and unregulated sale of antibiotics, long-standing patient expectations for antibiotics, low public health knowledge of AMR, and limited diagnostic testing capacity.^7,23–26^ A 2020 re-analysis of results from the GEMS study also estimated that there were approximately 12.6 inappropriate-treated diarrhea cases for each appropriately-treated case, with viruses (rotavirus, adenovirus, sapovirus) among the leading antibiotic-treated etiologies.^27^ Tools such as the App created for this study may provide clinicians with much-needed evidence to better identify patients with viral-only diarrhea and increase their confidence in following guidelines when antibiotic use is not indicated.

Used for a broad variety of applications in HICs, CDSSs have been beneficial for patient and health systems outcomes alike; CDSSs have been shown to improve antibiotic stewardship and reduce inappropriate prescription of antibiotics, reduce the use of high-cost computed tomography (CT) studies in emergency department patients, and improve glucose control in patients with diabetes, among many other examples.^28–30^ In the United States, a randomized controlled trial in rural communities in Utah and Idaho showed that an antibiotic stewardship CDSS run on a mobile device reduced overall antimicrobial use and improved appropriateness of antibiotic selection for respiratory tract infections.^31^ While a lack of research persists regarding CDSSs in LMICs, their use has risen recently due to the increasingly ubiquitous use of smartphones globally. Mobile CDSSs have been successfully used to improve hypertension control in India, and have been developed for neonatal care in Ghana, Kenya, and Uganda.^32–37^ In Bangladesh, electronic CDSSs have also been shown to improve WHO diarrhea guideline adherence including reducing non-indicated antibiotic use by 28.5% in children under five.^14^

While prior studies of CDSS targeting antibiotic use have focused on improving adherence to guidelines, our prediction tool provides the clinician with an estimation of etiology. Such information recognizes the expertise of the end-user and augments the available information that is used in the decision-making process for antibiotic use, potentially improving trust in the tool. In addition, entry of select clinical variables into the prediction tool may enhance clinician awareness of the relevance of such factors and further increase confidence in the use of the tool. A 2021 pre-implementation workshop among health professionals in Burkina Faso aimed at developing a CDSS for appropriate antimicrobial use in West Africa found that although the tool was expected to reduce antibiotic use, the lack of epidemiologic and microbiological data and limited availability of diagnostic tests were cited as anticipated barriers to use.^38^ Our App provides this additional information taking local epidemiologic data into account which may increase provider buy-in towards antibiotic stewardship efforts. Our study also provides an approach by which similar models could be developed for other infectious syndromes such as respiratory and febrile illness.

Importantly, all data collection for the App was conducted by nurses, the majority of whom had never used a mobile CDSS for clinical use previously. Nurses were able to confidently use the smartphone after a relatively brief training period and enter all necessary data into the tool without difficulty in high-volume clinical settings. The excellent reliability and agreement between nurses as well as between the App and the paper CRF reference demonstrate that accurate data collection is highly feasible in these clinical settings.

Our findings have several limitations. The study was conducted at study sites located in close proximity to sites from which the prediction model data were trained on. The prediction models may not therefore be generalizable to other locations, sites that have poor epidemiologic data on diarrhea etiologies, or locations that have initiated new vaccination campaigns (e.g. rotavirus). Additionally, approximately one-third of patients enrolled in this study did not have diarrhea etiology assigned using the predefined threshold AFe > 0.5 from the TAC testing data. The much higher rate of diarrhea etiology assignment in Bangladesh compared to Mali may be partially attributable to the study being conducted during rotavirus season in Bangladesh with rotavirus strongly associated with diarrhea and therefore easier to attribute. However, the predominance of rotavirus may limit the generalizability of this study’s findings. There is currently no existing gold standard for threshold of AFe that should be used to assign etiology and the threshold of 0.5 was set *a priori* based on expert consultation; however, the effect of using this cut-off has not yet been explored. A lower AFe threshold may still indicate likely etiology of diarrhea.

Despite these limitations, we have externally validated, in a prospective multicenter study, a smartphone-based clinical decision-support system that dynamically implements clinical prediction models based on real-time data streams. A clinical trial led by this study team is currently underway to evaluate the impact of this tool on antibiotic prescribing behaviors, and further research is needed to better understand how these tools could be best adapted for use by practicing clinicians in busy LMIC settings.

## Data Availability

The de-identified dataset is available upon reasonable request.

## Acknowledgements

The authors thank all study participants and the study staff of the icddr,b Dhaka Hospital and the health centers of Commune V and VI in Mali for their help and support. The authors also thank the development team at BeeHyv Software Solutions Pvt. Ltd. (Wilmington, DE; Hyderabad, India) who were instrumental in building the digital clinical decision support software used in this study.

## Supplemental File 1

List of Pathogen Targets for Taqman Array Card Testing

1. Adenovirus 40/41
2. *Aeromonas* spp.
3. Astrovirus
4. *Clostridium difficile*
5. *Campylobacteria jejuni/coli*
6. *Cryptosporidium spp*.
7. *Cyclospora spp*.
8. *Enterocytozoon bieneusi*
9. *Entamoeba histolytica*
10. *Encephalitozoon intestinalis*
11. Enteroaggregative *Escherichia coli* (EAEC)
12. Enteropathogenic *Escherichia coli* (EPEC)
13. Enterotoxigenic *Escherichia coli* (ETEC)
14. Shiga toxin-producing *Escherichia coli* (STEC)
15. *Giardia spp*.
16. *Helicobacter pylori*
17. *Isospora spp*.
18. Norovirus
19. *Plesiomonas spp*.
20. Rotavirus
21. *Salmonella* spp.
22. Sapovirus
23. *Shigella* spp.
24. *Vibrio cholerae*

## Supplemental File 2

Prediction density plot showing similar distributions of known etiology versus unknown etiologies.

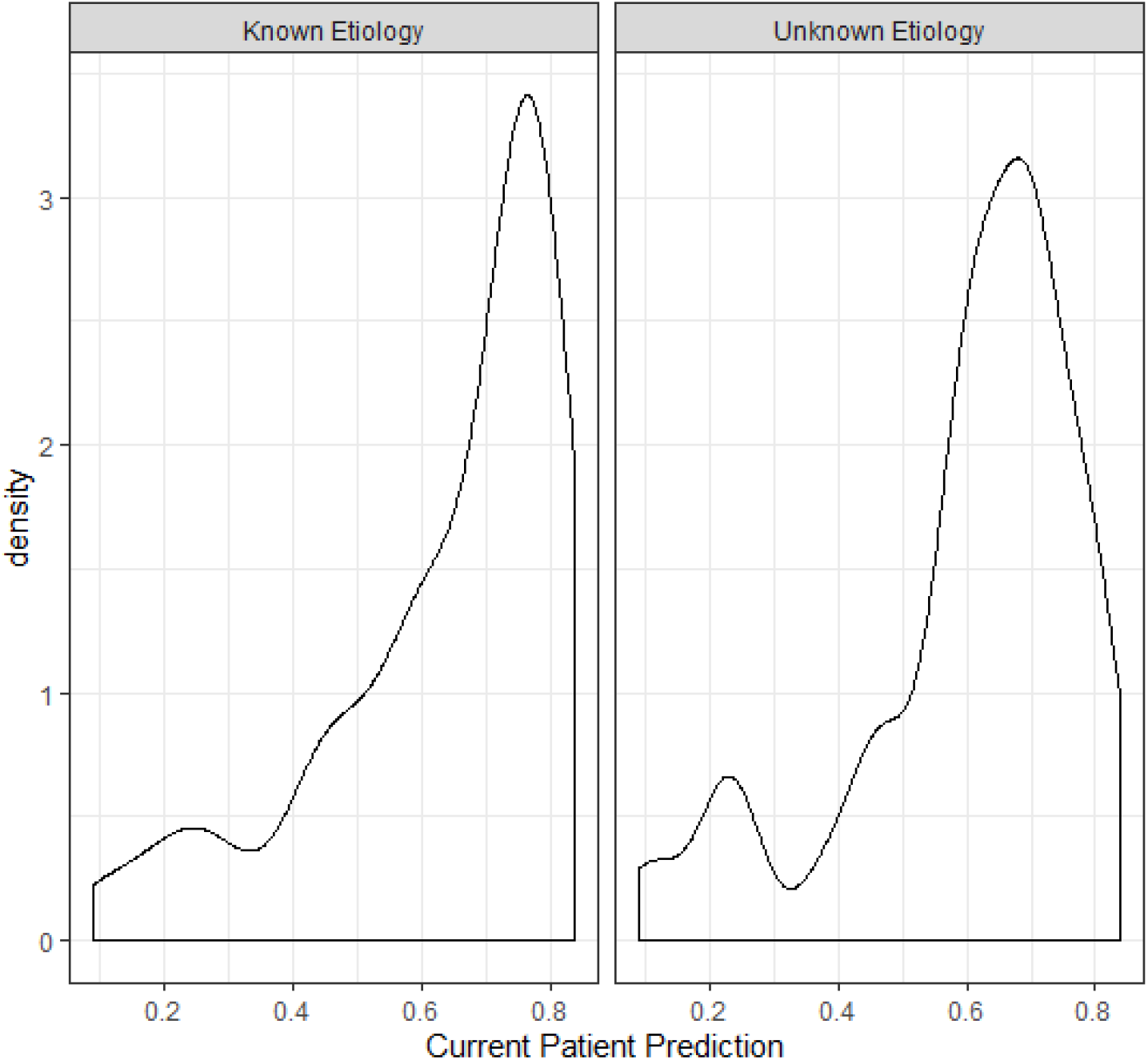

## Supplemental File 3

Descriptive data – demographics, predictors and viral-only outcome data from development dataset from GEMS

|  | Bangladesh<br>n(%)<br>701 | Mali<br>n(%)<br>432 |
| --- | --- | --- |
| Age (median, IQR), months | 18 (18) | 14 (14.3) |
| Sex | NA | NA |
| Male | 407 (58.1) | 230 (53.2) |
| Female | 294 (41.9) | 202 (46.8) |
| Bloody Stool Reported | NA | NA |
| Yes | 500 (71.3) | 45 (10.4) |
| No | 201 (28.7) | 387 (89.6) |
| Fever Reported | NA | NA |
| Yes | 579 (82.6) | 299 (69.2) |
| No | 122 (17.4) | 132 (30.6) |
| Vomiting Reported | NA | NA |
| Yes | 215 (30.7) | 172 (39.8) |
| No | 486 (69.3) | 260 (60.2) |
| Breastfeeding | NA | NA |
| Yes (Partial or Exclusive) | 565 (80.6) | 283 (65.5) |
| No | 136 (19.4) | 149 (34.5) |
| MUAC (median, IQR), cm | 14.03 (1.5) | 13.77 (1.9) |
|  | NA | NA |
| Viral-Only Etiology | 145 (20.7) | 122 (28.2) |
| Rotavirus-only etiology | 106 (15.1) | 88 (20.4) |
| *Abbreviations: IQR, interquartile range; cm, centimeter; MUAC, mid-upper arm circumference* |  |  |

## Supplemental File 4

Assessment of reliability and agreement between study nurses’ independent assessments of categorical predictor variables on case report forms.

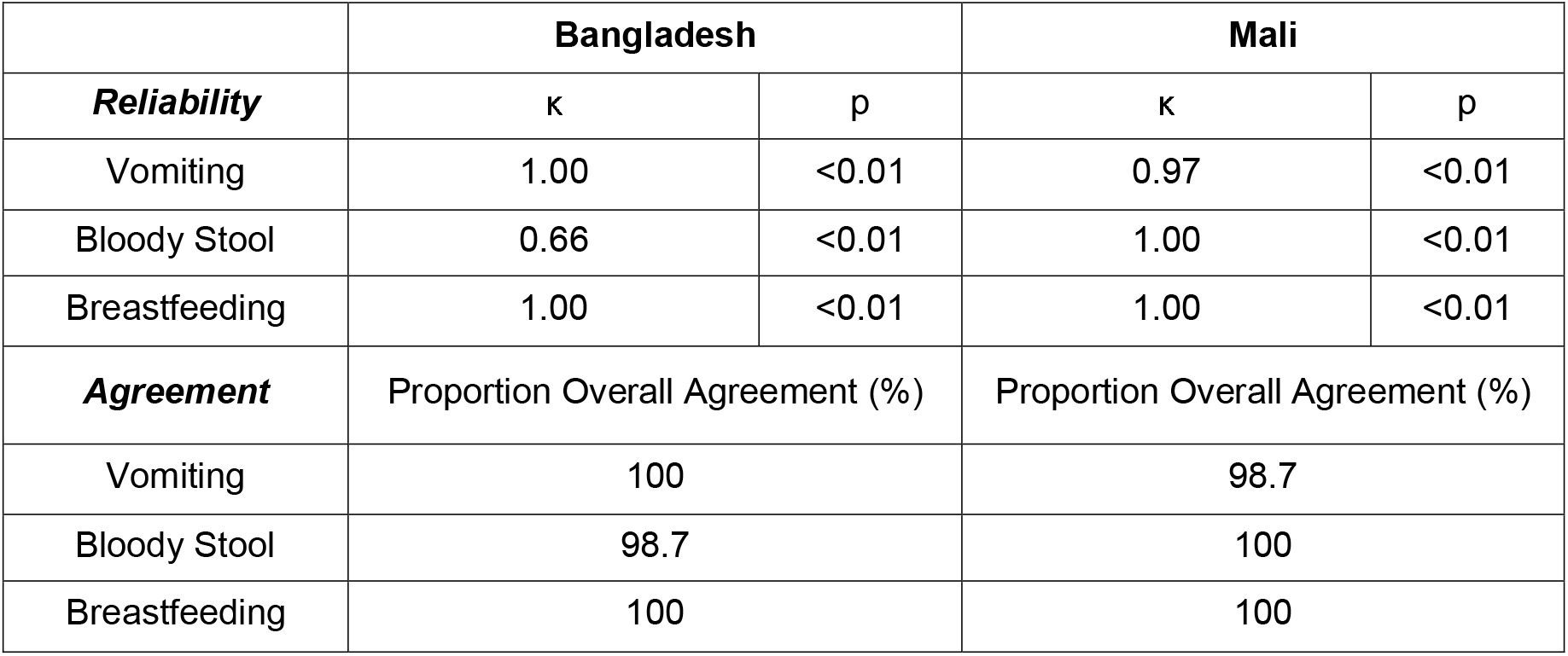

**Supplemental File 4.** Assessment of reliability and agreement between study nurses' independent assessments of categorical predictor variables on case report forms.
|  | **Bangladesh** |  | **Mali** |  |
| --- | --- | --- | --- | --- |
| ***Reliability*** | κ | p | κ | p |
| Vomiting | 1.00 | <0.01 | 0.97 | <0.01 |
| Bloody Stool | 0.66 | <0.01 | 1.00 | <0.01 |
| Breastfeeding | 1.00 | <0.01 | 1.00 | <0.01 |
| ***Agreement*** | Proportion Overall Agreement (%) |  | Proportion Overall Agreement (%) |  |
| Vomiting | 100 |  | 98.7 |  |
| Bloody Stool | 98.7 |  | 100 |  |
| Breastfeeding | 100 |  | 100 |  |

## Supplemental File 5

Agreement between study nurse recording on paper case record and input into App of categorical predictor variables.

|  | Bangladesh |  | Mali |  |
| --- | --- | --- | --- | --- |
| ***Reliability*** | $\kappa$ | p | $\kappa$ | p |
| Vomiting | 1.00 | <0.01 | 0.99 | <0.01 |
| Bloody Stool | 0.80 | <0.01 | 1.00 | <0.01 |
| Breastfeeding | NA<sup>a</sup> | NA<sup>a</sup> | 0.99 | <0.01 |
| ***Agreement*** | Proportion Overall Agreement (%) |  | Proportion Overall Agreement (%) |  |
| Vomiting | 100 |  | 99 |  |
| Bloody Stool | 99.3 |  | 100 |  |
| Breastfeeding | NA<sup>a</sup> |  | 99.7 |  |

<sup>a</sup> Data for breastfeeding did not sync successfully with the server in the Bangladesh study.

## Supplemental File 6

Bland-Altman plots showing agreement between data entered on case report form versus App for mid-upper arm circumference and calculated predicted viral-only etiology risk.

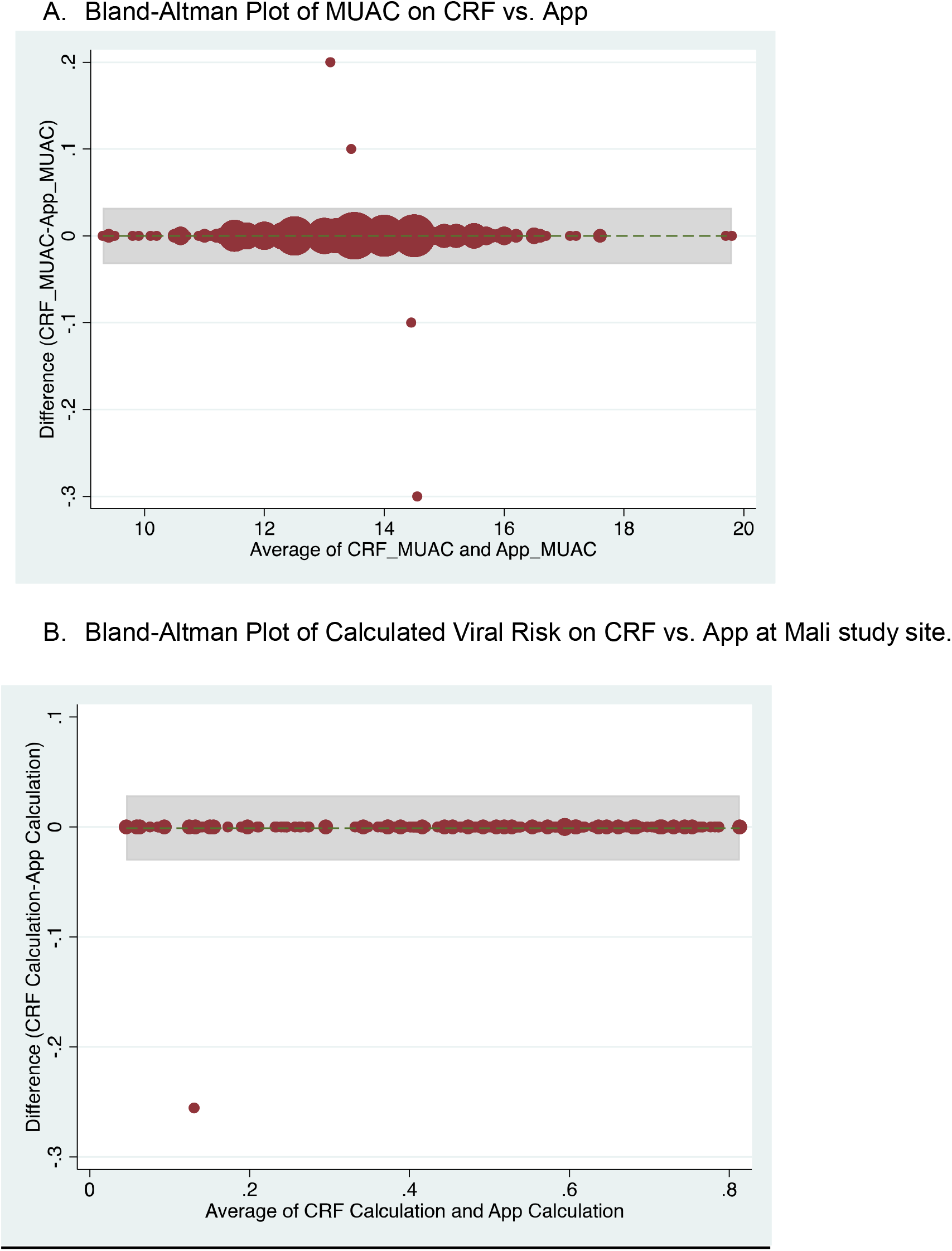

## Supplemental File 7

Congruence between the Application prediction of a patient’s viral etiology with the post-hoc prediction after adjusting for changing model development. Data shown are from the Mali study period alone because the software was updated between the Bangladesh and Mali study periods in response to engineering limitations.

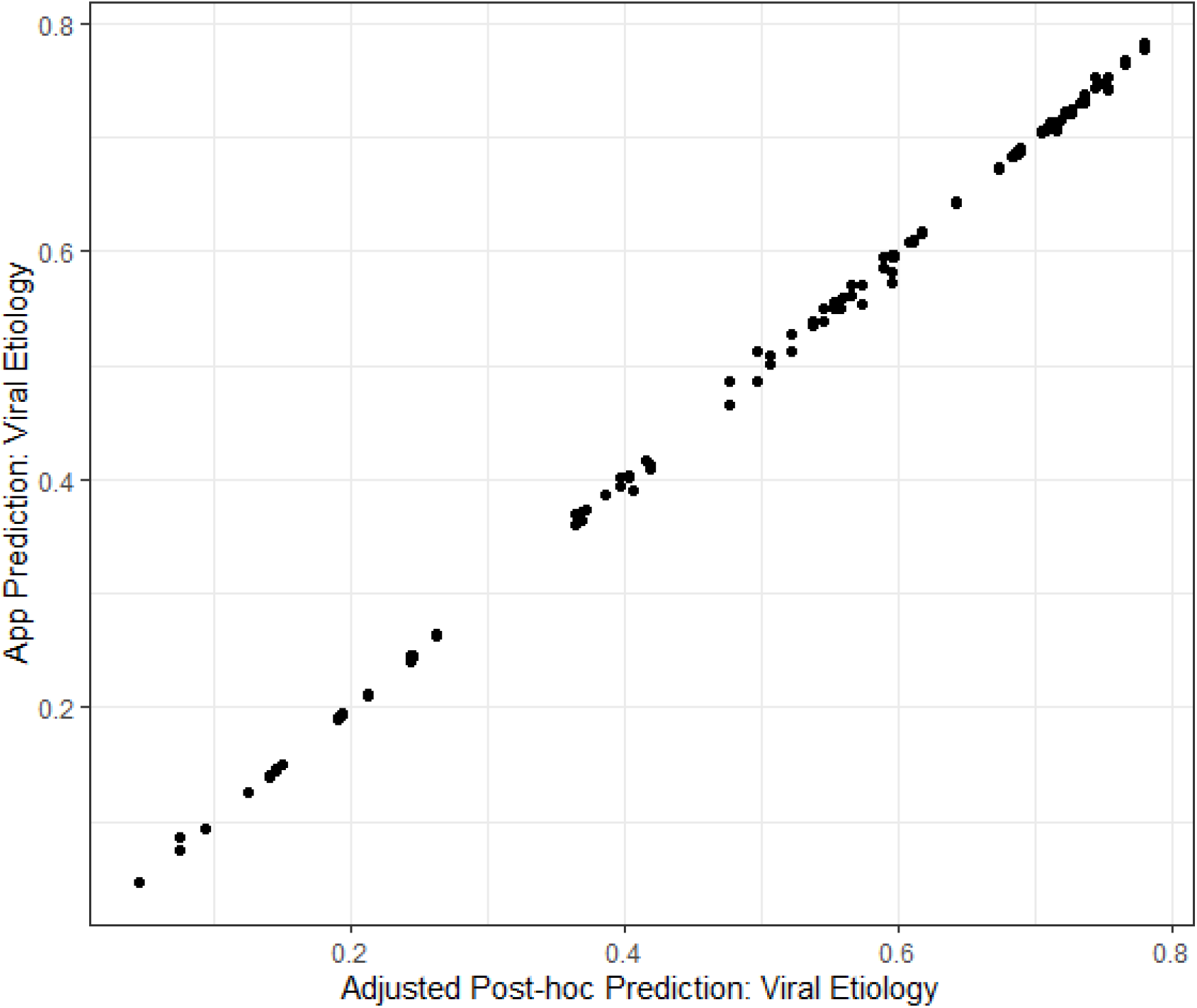

## Supplemental File 8

De-identified Dataset.

## References

1. James SL, Abate D, Abate KH, Abay SM, Abbafati C, Abbasi N, et al. Global, regional, and national incidence, prevalence, and years lived with disability for 354 Diseases and Injuries for 195 countries and territories, 1990-2017: A systematic analysis for the Global Burden of Disease Study 2017. Lancet. 2018 Nov 10;392(10159):1789–858.

2. Troeger C, Blacker BF, Khalil IA, Rao PC, Cao S, Zimsen SR, et al. Estimates of the global, regional, and national morbidity, mortality, and aetiologies of diarrhoea in 195 countries: a systematic analysis for the Global Burden of Disease Study 2016. Lancet Infect Dis. 2018 Nov 1;18(11):1211–28.

3. World Health Organization. THE TREATMENT OF DIARRHOEA A manual for physicians and other senior health workers. 2005.

4. Guarino A, Bruzzese E, Giannattasio A. Antibiotic treatment of acute gastroenteritis in children [Internet]. Vol. 7, F1000Research. Faculty of 1000 Ltd; 2018 [cited 2021 Apr 30]. Available from: /pmc/articles/PMC5814741/

5. Kotloff KL, Nataro JP, Blackwelder WC, Nasrin D, Farag TH, Panchalingam S, et al. Burden and aetiology of diarrhoeal disease in infants and young children in developing countries (the Global Enteric Multicenter Study, GEMS): a prospective, case-control study. www.thelancet.com [Internet]. 2013 [cited 2021 Apr 30];382. Available from: http://dx.doi.org/10.1016/

6. Platts-Mills JA, Liu J, Rogawski ET, Kabir F, Lertsethtakarn P, Siguas M, et al. Use of quantitative molecular diagnostic methods to assess the aetiology, burden, and clinical characteristics of diarrhoea in children in low-resource settings: a reanalysis of the MAL-ED cohort study. Lancet Glob Heal. 2018 Dec 1;6(12):e1309–18.

7. Bebell LM, Muiru AN. Antibiotic use and emerging resistance: How can resource-limited countries turn the tide? [Internet]. Vol. 9, Global Heart. Elsevier B.V.; 2014 [cited 2021 Apr 30]. p. 347–58. Available from: /pmc/articles/PMC4369554/

8. Kotloff KL. The Burden and Etiology of Diarrheal Illness in Developing Countries. Pediatr Clin North Am [Internet]. 2017 Aug 1 [cited 2019 Aug 24];64(4):799–814. Available from: http://www.ncbi.nlm.nih.gov/pubmed/28734511

9. Pavlinac PB, Denno DM, John-Stewart GC, Onchiri FM, Naulikha JM, Odundo EA, Hulseberg CE, Singa BO, Manhart LE, Walson JL. Failure of syndrome-based diarrhea management guidelines to detect shigella infections in kenyan children. J Pediatric Infect Dis Soc. 2016;5(4):366–74.

10. Bright TJ, Wong A, Dhurjati R, Bristow E, Bastian L, Coeytaux RR, Samsa G, Hasselblad V, Williams JW, Musty MD, Wing L, Kendrick AS, Sanders GD, Lobach D. Effect of Clinical Decision-Support Systems. Ann Intern Med [Internet]. 2012 Jul 3 [cited 2021 Apr 30];157(1):29. Available from: http://annals.org/article.aspx?doi=10.7326/0003-4819-157-1-201207030-00450

11. Tuon FF, Gasparetto J, Wollmann LC, de Moraes TP. Mobile health application to assist doctors in antibiotic prescription–an approach for antibiotic stewardship. Brazilian J Infect Dis. 2017;21(6):660–4.

12. Haque F, Ball RL, Khatun S, Ahmed M, Kache S, Chisti MJ, et al. Evaluation of a Smartphone Decision-Support Tool for Diarrheal Disease Management in a Resource-Limited Setting. PLoS Negl Trop Dis. 2017 Jan 19;11(1).

13. Brintz BJ, Haaland B, Howard J, Chao DL, Proctor JL, Khan AI, Ahmed SM, Keegan LT, Greene T, Keita AM. A modular approach to integrating multiple data sources into real-time clinical prediction for pediatric diarrhea. Elife. 2021;10:e63009.

14. Khan AI, Mack JA, Salimuzzaman M, Zion MI, Sujon H, Ball RL, et al. Electronic decision support and diarrhoeal disease guideline adherence (mHDM): a cluster randomized controlled trial. Lancet Digit Heal [Internet]. 2020 May 1 [cited 2021 May 17];2(5):e250–8. Available from: https://pubmed.ncbi.nlm.nih.gov/33328057/

15. Haque F, Ball RL, Khatun S, Ahmed M, Kache S, Chisti MJ, et al. Evaluation of a Smartphone Decision-Support Tool for Diarrheal Disease Management in a Resource-Limited Setting. PLoS Negl Trop Dis [Internet]. 2017 Jan 19 [cited 2021 May 17];11(1). Available from: https://pubmed.ncbi.nlm.nih.gov/28103233/

16. Liu J, Platts-Mills JA, Juma J, Kabir F, Nkeze J, Okoi C, et al. Use of quantitative molecular diagnostic methods to identify causes of diarrhoea in children: a reanalysis of the GEMS case-control study. Lancet. 2016 Sep 24;388(10051):1291–301.

17. Brintz BJ, Howard JI, Haaland B, Platts-Mills JA, Greene T, Levine AC, Nelson EJ, Pavia AT, Kotloff KL, Leung DT. Clinical predictors for etiology of acute diarrhea in children in resource-limited settings. PLoS Negl Trop Dis. 2020;14(10):e0008677.

18. Steyerberg EW, Vergouwe Y. Towards better clinical prediction models: seven steps for development and an ABCD for validation. Eur Heart J. 2014;35(29):1925–31.

19. Ahmed F, Farheen A, Ali I, Thakur M, Muzaffar A, Samina M. Management of Diarrhea in Under-fives at Home and Health Facilities in Kashmir. Int J Health Sci (Qassim) [Internet]. 2009 Jul [cited 2021 Apr 30];3(2):171–5. Available from: http://www.ncbi.nlm.nih.gov/pubmed/21475533

20. De T, Kondekar S, Rathi S. Hospital based prospective observational study to audit the prescription practices and outcomes of paediatric patients (6 months to 5 years age group) presenting with acute diarrhea. J Clin Diagnostic Res [Internet]. 2016 May 1 [cited 2021 Apr 30];10(5):SC01–5. Available from: /pmc/articles/PMC4948493/

21. Fink G, D’Acremont V, Leslie HH, Cohen J. Antibiotic exposure among children younger than 5 years in low-income and middle-income countries: a cross-sectional study of nationally representative facility-based and household-based surveys. Lancet Infect Dis. 2020 Feb 1;20(2):179–87.

22. Tsige AG, Nedi T, Bacha T. <p>Assessment of the Management of Diarrhoea Among Children Under Five in Addis Ababa, Ethiopia</p>. Pediatr Heal Med Ther [Internet]. 2020 May [cited 2021 Apr 30];Volume 11:135–43. Available from: /pmc/articles/PMC7213891/

23. Vila J, Pal T. Update on Antibacterial Resistance in Low-Income Countries: Factors Favoring the Emergence of Resistance. Vol. 4, The Open Infectious Diseases Journal. 2010.

24. Okeke IN, Laxminarayan R, Bhutta ZA, Duse AG, Jenkins P, O’Brien TF, Pablos-Mendez A, Klugman KP. Antimicrobial resistance in developing countries. Part I: recent trends and current status. Lancet Infect Dis. 2005;5(8):481–93.

25. World Health Organization. Global action plan on antimicrobial resistance. WHO [Internet]. 2017 [cited 2019 Aug 20]; Available from: https://www.who.int/antimicrobial-resistance/publications/global-action-plan/en/

26. Klein EY, Van Boeckel TP, Martinez EM, Pant S, Gandra S, Levin SA, Goossens H, Laxminarayan R. Global increase and geographic convergence in antibiotic consumption between 2000 and 2015. Proc Natl Acad Sci U S A [Internet]. 2018 Apr 10 [cited 2021 Apr 30];115(15):E3463–70. Available from: www.pnas.org/cgi/doi/10.1073/pnas.1717295115

27. Lewnard JA, Rogawski McQuade ET, Platts-Mills JA, Kotloff KL, Laxminarayan R. Incidence and etiology of clinically-attended, antibiotic-treated diarrhea among children under five years of age in low- and middle-income countries: Evidence from the Global Enteric Multicenter Study. Freeman MC, editor. PLoS Negl Trop Dis [Internet]. 2020 Aug 10 [cited 2021 Apr 30];14(8):e0008520. Available from: https://dx.plos.org/10.1371/journal.pntd.0008520

28. Sintchenko V, Coiera E, Gilbert GL. Decision support systems for antibiotic prescribing. Curr Opin Infect Dis [Internet]. 2008 Dec [cited 2021 Apr 30];21(6):573–9. Available from: http://journals.lww.com/00001432-200812000-00002

29. Bookman K, West D, Ginde A, Wiler J, McIntyre R, Hammes A, Carlson N, Steinbruner D, Solley M, Zane R. Embedded clinical decision support in electronic health record decreases use of high-cost imaging in the emergency department: Emb ED study. Acad Emerg Med. 2017;24(7):839–45.

30. Carracedo-Martinez E, Gonzalez-Gonzalez C, Teixeira-Rodrigues A, Prego-Dominguez J, Takkouche B, Herdeiro MT, Figueiras A, Group GPR. Computerized clinical decision support systems and antibiotic prescribing: a systematic review and meta-analysis. Clin Ther. 2019;41(3):552–81.

31. Samore MH, Bateman K, Alder SC, Hannah E, Donnelly S, Stoddard GJ, Haddadin B, Rubin MA, Williamson J, Stults B, Rupper R, Stevenson K. Clinical Decision Support and Appropriateness of Antimicrobial PrescribingA Randomized Trial. JAMA [Internet]. 2005 Nov 9;294(18):2305–14. Available from: https://doi.org/10.1001/jama.294.18.2305

32. Watson HA, Tribe RM, Shennan AH. The Role of Medical Smartphone Apps in Clinical Decision-Support: A Literature Review. Artif Intell Med. 2019;101707.

33. Anchala R, Kaptoge S, Pant H, Emanuele M;, Angelantonio D, Franco OH, Prabhakaran; D. Evaluation of Effectiveness and Cost-Effectiveness of a Clinical Decision Support System in Managing Hypertension in Resource Constrained Primary Health Care Settings: Results From a Cluster Randomized Trial. [cited 2021 Apr 30]; Available from: http://www.ctri.nic.in.

34. Muhindo M, Bress J, Kalanda R, Armas J, Danziger E, Kamya MR, Butler LM, Ruel T. Implementation of a Newborn Clinical Decision Support Software (NoviGuide) in a Rural District Hospital in Eastern Uganda: Feasibility and Acceptability Study. JMIR mHealth uHealth. 2021;9(2):e23737.

35. Bucher SL, Rajapuri A, Ravindran R, Rukunga J, Horan K, Esamai F, Purkayastha S. The Essential Care For Every Baby Digital Action Plan: Design And Usability Testing Of A Mobile Phone-Based Newborn Care Decision Support Tool In Kenya. Am Acad Pediatrics; 2021.

36. Amoakoh HB, Klipstein-Grobusch K, Amoakoh-Coleman M, Agyepong IA, Kayode GA, Sarpong C, Grobbee DE, Ansah EK. The effect of a clinical decision-making mHealth support system on maternal and neonatal mortality and morbidity in Ghana: study protocol for a cluster randomized controlled trial. Trials. 2017;18(1):1–11.

37. Bilal S, Nelson E, Meisner L, Alam M, Amin S Al, Ashenafi Y, Teegala S, Khan AF, Alam N, Levine A. Evaluation of standard and mobile health-supported clinical diagnostic tools for assessing dehydration in patients with diarrhea in rural Bangladesh. Am J Trop Med Hyg. 2018;99(1):171–9.

38. Peiffer-Smadja N, Poda A, Ouedraogo AS, Guiard-Schmid JB, Delory T, Le Bel J, Bouvet E, Lariven S, Jeanmougin P, Ahmad R, Lescure FX. Paving the way for the implementation of a decision support system for antibiotic prescribing in primary care in west Africa: Preimplementation and co-design workshop with physicians. J Med Internet Res [Internet]. 2020 Jul 1 [cited 2021 Apr 30];22(7):e17940. Available from: https://www.jmir.org/2020/7/e17940

